# COVID-19 European regional tracker

**DOI:** 10.1101/2021.02.15.21251788

**Authors:** Asjad Naqvi

**Affiliations:** Advancing Systems Analysis (ASA), International Institute for Applied Systems Analysis (IIASA), Laxenburg, 2361, Austria; Department of Socioeconomics, Vienna University of Economics and Business (WU), Vienna, 1020, Austria

**Author notes:** Corresponding author: Asjad Naqvi.

## Abstract

This Tracker presents data on daily COVID-19 cases at the sub-national level for 26 European countries from January 2020 till present. Country-level data sources are identified and processed to form a homogenized panel at the NUTS 3 or NUTS 2 level, the two lowest standardized administrative units of Europe. The strengths and weaknesses of each country dataset are discussed in detail. The raw data, spatial layers, the code, and the final homogenized files are provided in an online repository for replication. The data highlights the spatial distribution of cases both within and across countries that can be utilized for a disaggregated analysis on the impacts of the pandemic. The Tracker is updated monthly to expand its coverage.

## Background & Summary

The COVID-19 European Regional Tracker (henceforth referred to as the Tracker)^1^ collates sub-national information for cumulative and daily reported COVID-19 cases for 26 countries in Europe starting from 15th January 2020 till present. Data sources of each country are discussed in detail, including their strengths and weaknesses, and the raw country-level files are provided in an online repository. Additional effort has been put into homogenizing this data at the NUTS 3 level for all countries in the Tracker. For two countries, Poland and Greece, the complete data is only available at the NUTS 2 level. NUTS stands for Nomenclature of Territorial Units for Statistics and represent standardized administrative units defined by the European Commission for reporting various regional statistics on Europe.^2^ The reason for creating a homogenized dataset at the NUTS level is to allow the Tracker to be easily merged with other regional datasets. For example, Eurostat, the official statistical agency of the European Commission^3^, provides a host of economic, demographic, health and other indicators. In addition to this, regional data from national-level statistical agencies in European countries mostly utilize NUTS definitions. Furthermore, several recent global-level datasets, like Google mobility trends^4^ or the Facebook Social Connectedness Index^5^, have structured their regional data around NUTS regions for European countries.

COVID-19 cases exploded in Europe around early March 2020. At the center of this spread were the regions of North Italy^6^ and the ski resort of Ishgl in the western part of Austria.^7^ From this point onward, the virus quickly spread across the European continent, resulting in a rapid increase in cases and deaths, and alarming governments that implemented stringent lockdown measures including border closures, reductions in mobility, to shutting down the economy.^8^ While the virus was mostly contained during the summer of 2020, a resurgence in cases in the Fall of 2020 resulted in a massive second wave that completely overshadowed the first wave in terms of cases and deaths. In order to contain the virus, a second round of lockdown measures were put in place around October 2020.^9^ In early 2021, the detection of more infectious virus variants, and a slower-than-expected vaccine roll-out in Europe prompted countries to further extend their lockdown measures. As Figure 1 shows that no other continent has been impact by the virus as much as Europe, which still has a large share in global cases and deaths.

**Figure 1.**
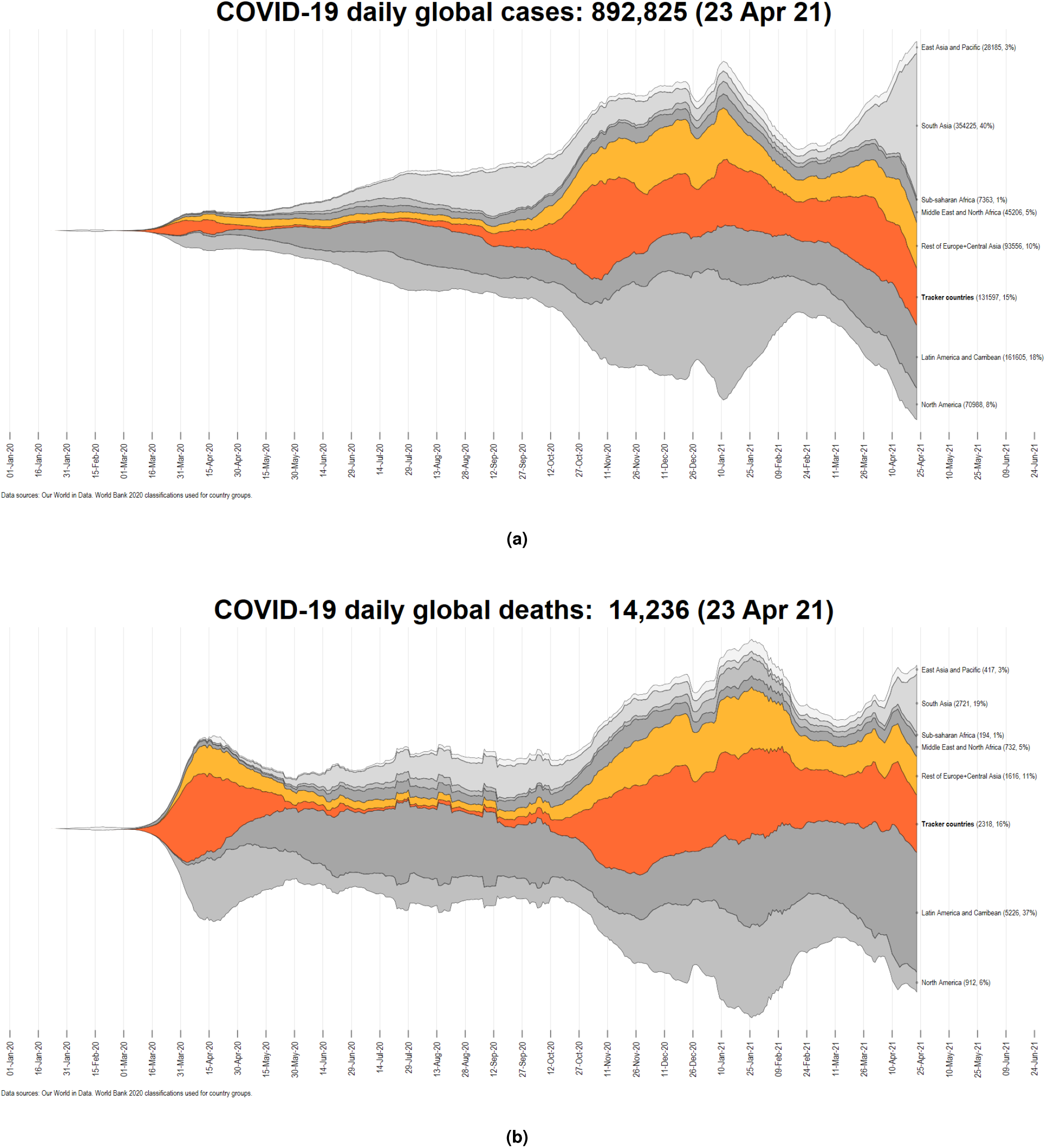
Global distribution of COVID-19 cases and deaths. Countries in the tracker are highlighted in dark orange, while the rest of Europe is shown in a light orange color.

A unique feature of the COVID-19 pandemic is the amount of knowledge and data that is constantly being generated to understand how this event unfolds. For a high-income region like Europe, the quality of information that is made available on a daily basis is exceptionally high as compared to the rest of the world. Furthermore, several innovative datasets have appeared since the start of the pandemic that collect unique COVID-19 related information. For example, the Oxford COVID-19 Government Response Tracker^10^ and the Complexity Science Hub (CSH) Tracker^11^ evaluate daily policy changes for a host of indicators for all countries of Europe, and their coverage extends to the rest of the world as well. Our World in Data (OWID), a website that curates various global data sets, has produced a tracker on COVID-19 tests performed^12^ and is currently leading the efforts to document the vaccine roll-out across the globe. Google has released information how mobility is evolving over time^4^ and Facebook has released data on how connected regions are with each other.^5^ Various other data sources can be viewed on the Oxford COVID-19 Super-Tracker website (https://supertracker.spi.ox.ac.uk/policy-trackers/) that has catalogued over a hundred different projects.^13^

In Europe, almost all countries provide information through interactive dashboards, maps, and data visualizations. Before October 2020, COVID-19 information for European countries was collected daily by the European Centre for Disease Prevention and Control (ECDC), and NUTS 2 level maps were regularly released to track regional trends.^14^ In November 2020, ECDC decided to stop daily updates and switched to aggregated weekly reporting interval of NUTS-2 or higher level data. Since March 2021, country-level daily data is again available, but currently the weekly situation reports (https://www.ecdc.europa.eu/en/geographical-distribution-2019-ncov-cases) are the main focus.^15^ Since ECDC was the official source of COVID-19 related information for Europe, the reduction in the frequency of data availability resulted in a major data gap for tracking how the virus is evolving in the continent. As a result, data aggregation websites like Our World in Data (OWID) (https://ourworldindata.org/coronavirus) switched to John Hopkins University COVID-19 database (https://coronavirus.jhu.edu/map.html) to maintain the daily reporting frequency.^16^ On a positive side, in the summer of 2020, almost all European countries increased their efforts to display and share regional data at a daily frequency on various official online platforms.

The aim of this Tracker is to identify, collect, and collate various official regional dataset for European countries. This tracker, while providing raw regional-level data, also combines and homogenizes the data at the NUTS 3 or NUTS 2 level. This homogenized dataset allows us to explore how the virus spreads in terms of cumulative cases, daily cases, and cases per capita in Europe at a daily resolution. In this Tracker, country-level data sources and their strengths and weaknesses are discussed in detail. Country-wise regional data, and the Stata code that compiles the data is updated on GitHub (https://github.com/asjadnaqvi/COVID19-European-Regional-Tracker) approximately every four weeks.^1^ Raw and homogenized data files are also provided in the common CSV format which allows users to import the data or replicate the code in other software languages as well.

## Methods

Figure 2 shows the workflow for the Tracker. In the first step, each country’s data source is identified together with its spatial unit of analysis. The source can either be official or scraped data depending on how open the country is about sharing it’s data especially in a machine-readable format. These raw files are saved as part of the Tracker to allow users access to the original information. Data on cases is extracted from all the raw files since this variable is the lowest common denominator that exists for all the countries. The raw data also contains additional variables like, deaths, tests performed, hospitalization rates, and vaccination rates. Sometimes, detailed breakdowns by age groups and gender is also available. These additional variables can be easily extracted from the raw data as well. In the second step, the raw files are homogenized to NUTS 3 2016 boundaries by using official EU correspondence tables. If these are not available, a crosswalk between different administrative boundaries and NUTS 2016 layers is extracted by through a spatial overlay in QGIS (https://qgis.org/en/site/), an open-source mapping software. If NUTS 3 data is not available then NUTS 2 boundaries are used, for example in the case of Greece and Poland.

**Figure 2.**
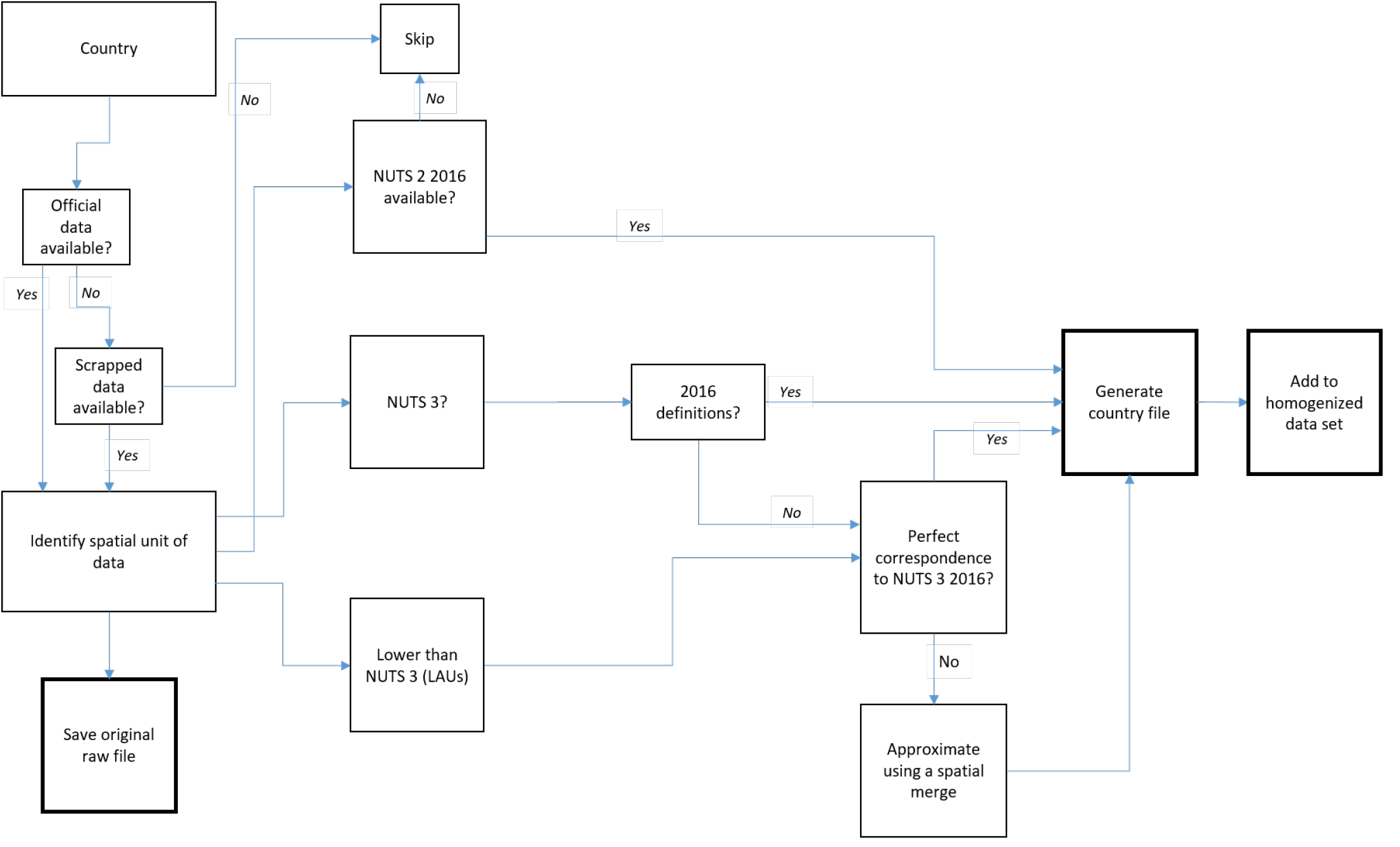
Workflow for the Tracker

Countries in Europe define regions differently, and therefore, making data homogeneous is a challenging task. For consistency, Eurostat, the official agency of the European Union (EU), uses standardized units called Nomenclature of Territorial Units for Statistics or NUTS (https://ec.europa.eu/eurostat/web/nuts/history).^2^ NUTS 0 represent countries, NUTS 1 are provinces, NUTS 2 broadly represent districts, and NUTS 3 are broadly defined as municipalities or other sub-divisions of districts. Each country independently defines its own administrative units that are mapped onto NUTS regions. Regions below NUTS 3, are referred to as Local Administrative Units (LAUs) (https://ec.europa.eu/eurostat/web/nuts/local-administrative-units) that were formerly NUTS 4 and NUTS 5 tiers.^17^ Most countries provide data at the LAU level.

Table 1 summarizes the regional classifications of countries currently in the Tracker. The table shows the mapping of country-level regions together with the number of administrative units within that regional classification in brackets. The administrative unit at which the data is available is highlighted in bold. For most countries this is at either NUTS 3 level or lower. Two countries, Poland and Greece, are mapped at the NUTS 2 level since data is only available at this resolution for the whole duration of the tracker. United Kingdom (UK) is dealt with as four separate countries: England, North Ireland, Scotland, and Wales. This is because each country has it’s own COVID-19 dashboard and the centralized COVID-19 database for the UK has put restrictions on bulk data access (https://coronavirus.data.gov.uk/details/download). An additional challenge that was faced in creating this Tracker was to navigate the different websites of individual countries that are usually in their native languages. Hence, there were significant time investment costs in translating the websites, and identifying the correct files and the variables.

**Table 1.**
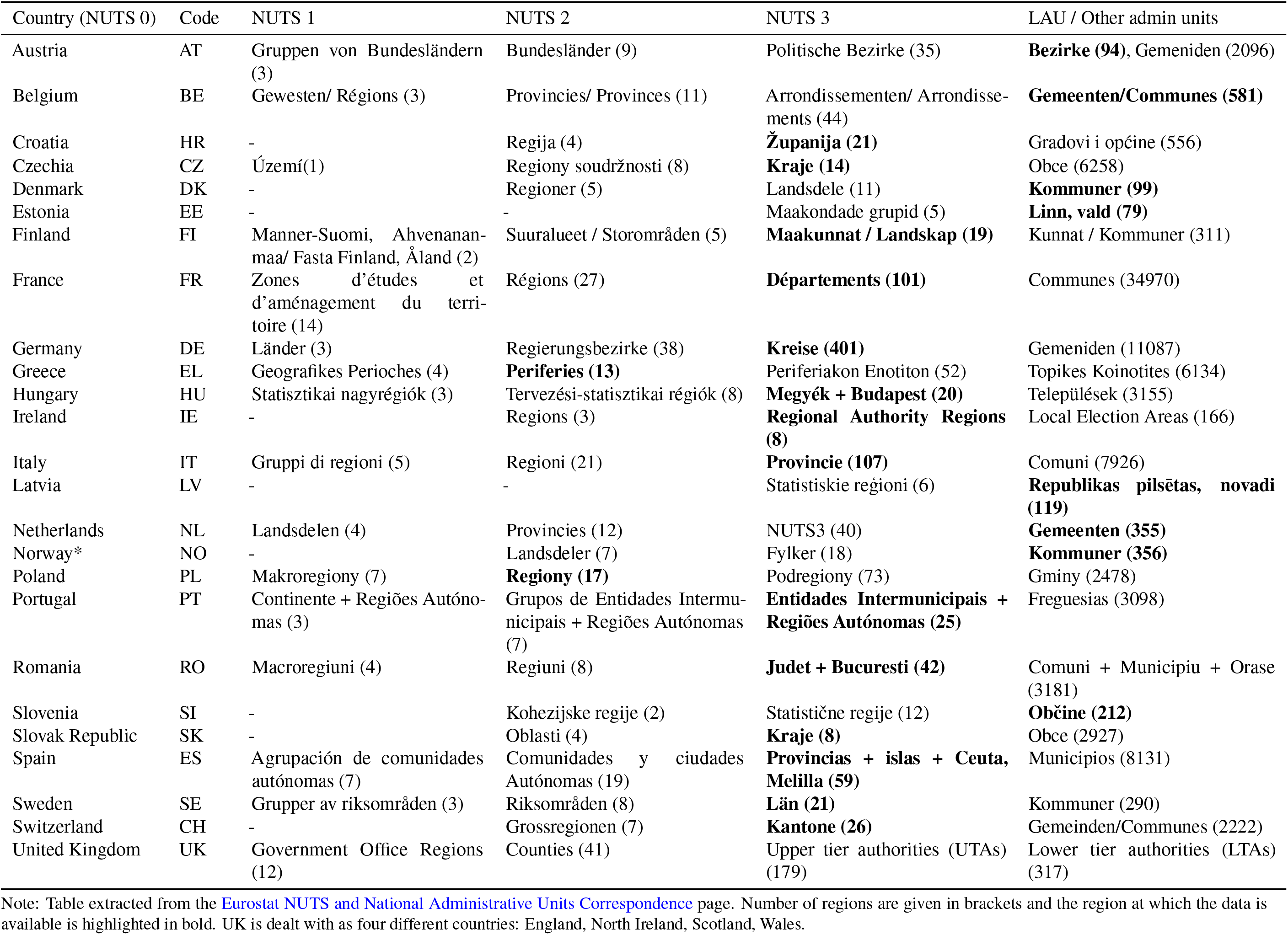
European regional classifications

On the data homogenization side, two main challenges exist with the regions-to-NUTS mapping. First, NUTS are re- classified every few years (2003, 2006, 2010, 2013, 2016, 2021), mostly due to demographic changes that can result in boundary shifts, and splitting or merging of regions. Since the epidemic started in 2020, 2016 definitions were in place and therefore the tracker also homogenizes the data to NUTS 2016 boundaries. NUTS 2016 are also the definitions currently used by Eurostat for regional data since currently only 2020 data is available for majority of the indicators. The boundary data is provided by Eurostat’s GISCO or the Geographic Information System of the Commission.^18^ As of 1st January 2021, the NUTS 2021 definitions have come into effect, and countries might switch to reporting on the new boundaries depending on how long the pandemic lasts. While most of the regions remain unchanged, minor shifts in boundaries can result in imperfect matching for some regions. A good example of this is Italy which already started reporting data at the NUTS 2021 boundaries in 2020. These bottom-up aggregation errors are highlighted and discussed in the Technical Validation section together with the extent of the errors. Regardless of the minor matching issues in the homogenized dataset, raw data is also available in case the users prefer to use the original source of information.

Second, some countries use different types of administrative divisions for regional tracking of COVID-19 cases that do not have an official correspondence to NUTS classifications. For example, Finland uses Hospital Districts, Greece uses Prefectures, Norway uses Kommunes, and the UK reports information at the Local Authority Districts (LADs). This issue is resolved by overlaying the boundaries of these administrative regions with NUTS 2016 boundaries to generate a spatial region-to-NUTS mapping. For Greece, Norway, and the UK the regions map perfectly to NUTS boundaries while for Finland, small errors persist in some regions. These are also highlighted and discussed in the Technical Validation section.

### Data Records

The data for the tracker has been made publicly available on Zenodo under the Creative Commons Attribution 4.0 International Licence (CC-BY). The latest version of the repository can be accessed using a stable Zenodo DOI: https://doi.org/10.5281/zenodo.4244878.^1^ The Tracker data in this paper shows the evolution of cases in European regions from January 2020 till April 2021. Since the COVID-19 virus started spreading in Europe around March 2020, the panel of the country-regions- date is mostly complete from April 2020 onward. Figure 3 summarizes the exact date range for each country. The GitHub repository (https://github.com/asjadnaqvi/COVID19-European-Regional-Tracker) is updated once a month to expand its temporal coverage, and fix errors and bugs if any. But if users want a higher update frequency, all the code and links to the online repositories are also provided on the Tracker page and can be utilized for more frequent updates.

**Figure 3.**
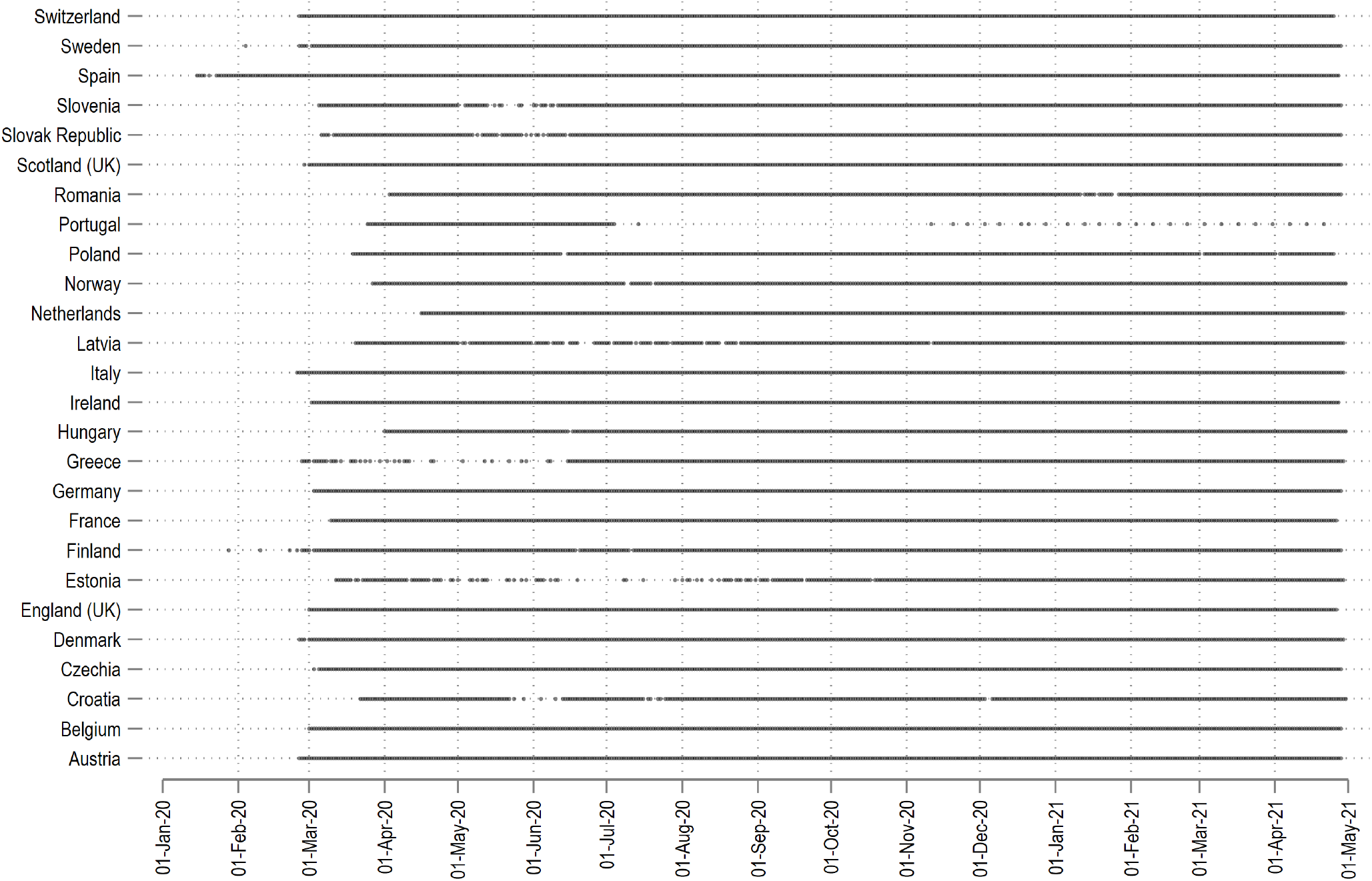
Data range of countries in the Tracker.

For each country, two sets of data files are provided. The first set contains the raw data files downloaded from various sources. These files include the records as they exist in the original data including the information on the spatial unit at which the information is released. Most countries provided data at a administrative units below NUTS 3 (see Table 1). Furthermore, most of the raw files contain more information than in the final data set homogenized at the NUTS level which contains cumulative and daily cases information, the baseline variables that exist in all country files. Additional variables in the raw data, for example, include deaths, recovered, tested, hospitalized, and vaccinated. Some countries also provide age and gender-wise breakdowns. Therefore, users of this Tracker can go deeper with their analysis by compiling a finer resolution and more detailed dataset for analysis. The second set are the processed country files that map and convert the raw data into homogenized datasets at the NUTS level. This mapping is done using Stata scripts or dofiles, which which process the data and homogenize the region-to-NUTS mapping. If Stata is not available, the dofiles can be viewed with any generic text editor for the code structure. The code structure is straightforward to read and can be easily converted to other programming languages.

The files on GitHub (https://github.com/asjadnaqvi/COVID19-European-Regional-Tracker) and Zenodo^1^ are sorted in the folder structure shown in Box 1:

#### Box 1

Folder structure

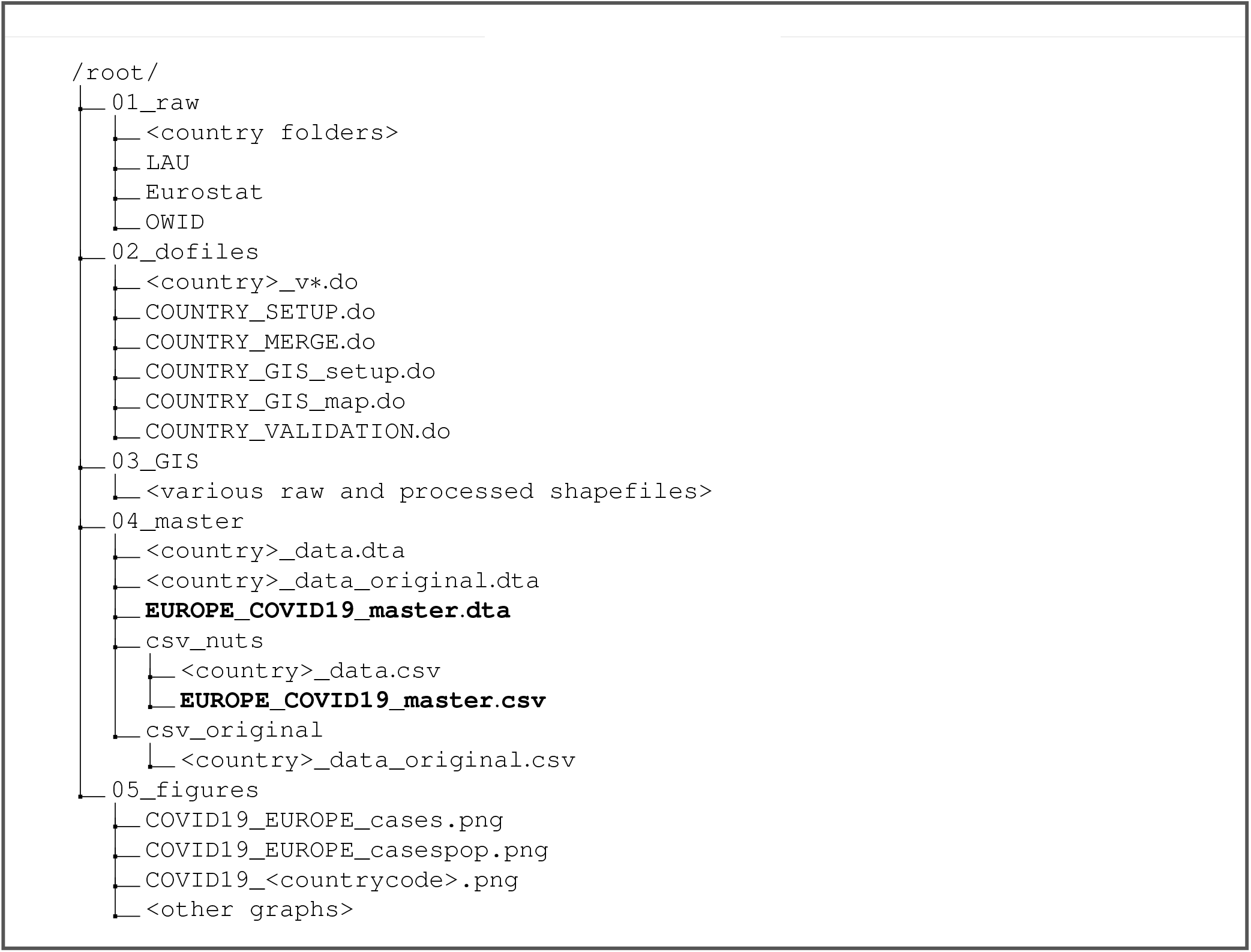

The folders are described as follows:

- **01_raw** contains miscellaneous country-level files. Each country has its own sub-folder with all the files necessary to generate a clean version of the raw and homogenized data. These folders also include various files that help map region identifiers to NUTS classifications. The raw data itself is saved in a Stata .dta format and the generic .csv file format in the 04_master folder. The LAU folder contains the LAU 2019-to-NUTS 2016 correspondence file^2^ from which several country-level files are extracted where necessary. The Eurostat folder contains the regional population file in Stata format extracted from the demo_r_pjangrp3 data table (https://ec.europa.eu/eurostat/databrowser/view/demo_r_pjangrp3/). For the Tracker, the 2020 regional population values are used except for UK, where due to Brexit regional data information is only available till 2019. OWID folder contains data from the Our World in Data^12^ website which is used for validating the Tracker.
- **02_dofiles** contains the Stata scripts called dofiles for each country, and five additional dofiles that compile, merge, map, and validate the data (see the Code Availability section for details). The dofiles are also version controlled (_v1, _v2,
- _v3 etc.) to track changes in data sources and data structures. Only the latest dofile for each country is uploaded to the repository. Each country dofile saves the raw data, and processes the raw data to created the homogenized file, both of which are saved in the 04_master folder. See the Code Availability section for details on how to run these files.
- **03_GIS** contains the raw and processed GIS files. The raw NUTS 0 to NUTS 3 2016 shapefiles are downloaded from Eurostat’s GISCO website^18^ and processed using the COUNTRY_GIS_setup.do in the **02_dofiles** folder.
- **04_master** contains the raw and the homogenized country files, and the final dataset **EUROPE_COVID19_master.dta**. The .csv versions of the the raw and homogenized files are saved in the csv_original and csv_nuts folders respectively.
- **05_figures** contains the maps and figures generated from various dofiles.

The main data file “**COVID19_master.dta**” and its .csv version are provided in the 04_master folder. The master files contain the following variables with their description given in brackets:

|  |  |
| --- | --- |
| nuts0_id | (country code) |
| nuts2_id | (for Greece and Poland only) |
| nuts3_id |  |
| nuts_id | (combined NUTS 2 and NUTS 3) |
| date |  |
| cases | (cumulative cases) |
| population | (extracted from Eurostat population file) |
| cases_daily | (daily new cases) |
| cases_daily_pop | (daily new cases per 10,000 population) |

Table 2 summarizes the individual country sources of raw data. For the 26 countries currently in the Tracker, the table lists their respective departments that collect and disseminate COVID-19 data, links to official COVID-19 dashboards, and links to data repositories that are used to pull the data for this Tracker. The very precise paths are given in the Stata dofile of each country and if Stata is not available, the they can be viewed in any text editor. Each country’s raw data files are also saved in the 04_master folder with the suffix _original.dta and _original.csv. Please note that these links are also subject to change and for the latest information check the information on GitHub.^1^

**Table 2.**
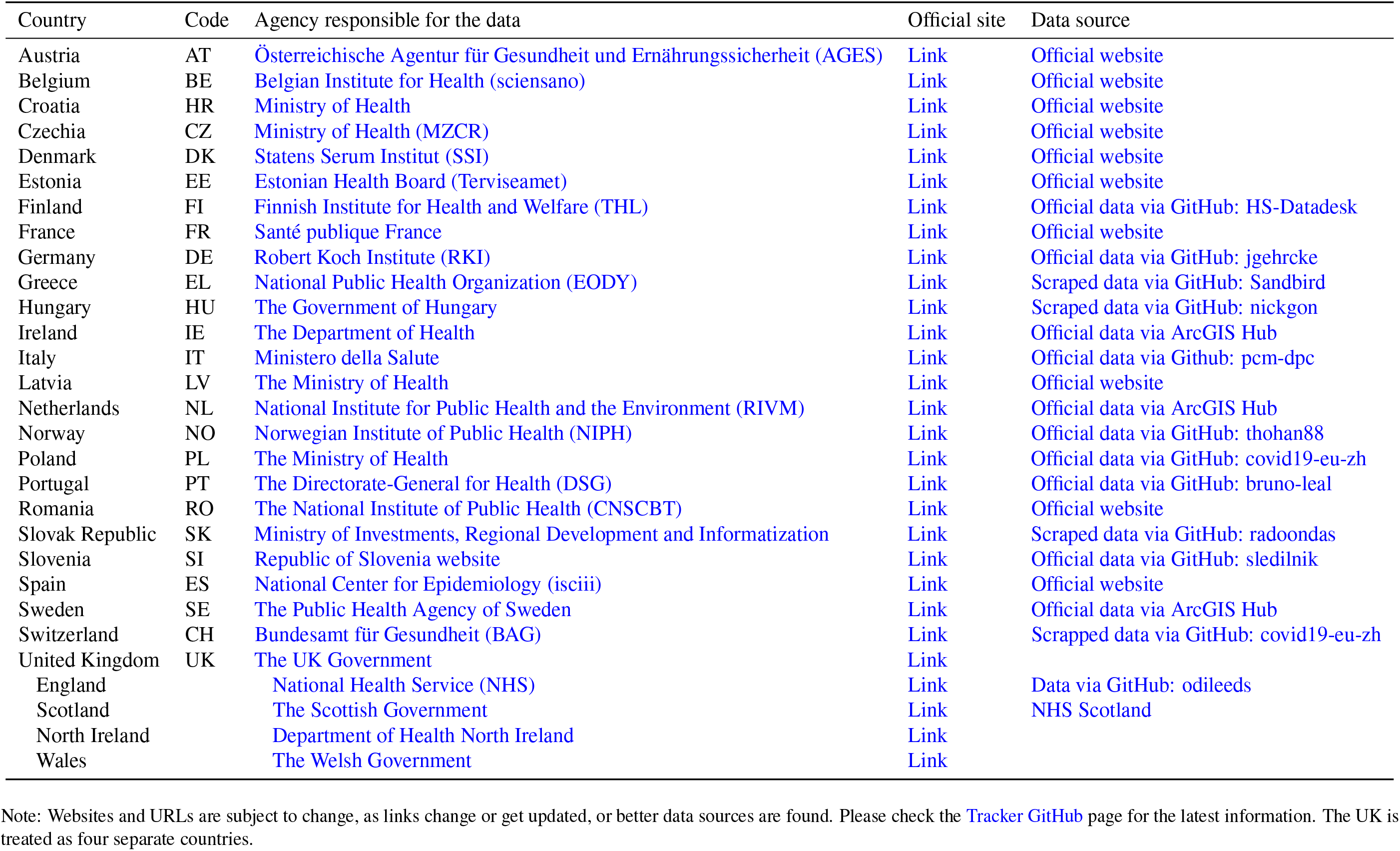
Country-specific data sources

### Data for individual countries

Data for the 26 countries in the Tracker is briefly discussed below including the challenges:

- **Austria**: Austria currently provides daily updates at the district (Bezirk) level, a tier below NUTS 3. The data can be downloaded directly from the official website as a zipped file from which the relevant file is extracted and processed.
- **Belgium**: Belgium provides daily updates and the data file can be read directly from the website. Regions with cases 5 or below are labeled as ≤ 5 for privacy reasons. As a judgement call, to keep the variable numeric, these values have been replaced with 1. Users can refer to the original data or change the data scripts if a different way of dealing with ranges is preferred.
- **Croatia**: Croatia provides official data in a JSON format which is read directly from the website.
- **Czechia**: Czechia data is imported directly from the official website for processing.
- **Denmark**: Denmark releases a daily zipped file which is downloaded manually from the website. The zipped file contains several data files from which the one containing information on cases is used.
- **Estonia**: Estonia officially releases the data on COVID-19 but in order to maintain privacy, data is only provided in ranges of 10s, for example, 10-20, 20-30, 100-110, etc. In order to make this variable numeric, the mid-points of these ranges are taken, and aggregated at the NUTS 3 level. Using mid-points is a judgement call to make the cases variable numeric. The raw data file contains the original information which can be processed differently if required.
- **Finland**: The data for Finland is taken from a GitHub repository that scrapes the data and provides it in a JSON format. This is read and parsed directly in Stata. This data is at the Hospital District level which does not perfectly correspond to NUTS 3 boundaries and are approximated using a spatial merge. See Technical validation section for detailed notes.
- **France**: France provides a comprehensive range of COVID-19 related indicators on its official website. France also switched methodologies for improving the data quality in May 2020. Old data is available online and has been merged with the current data but the quality is poor. Data of territories outside of mainland Europe have been dropped from the Tracker. The file is downloaded manually from the website for processing.
- **Germany**: Germany’s regional data is provided by the Robert Koch Institute (RKI) and has been of high quality data since the early days of the pandemic. Several repositories of this data exist on GitHub, of which, one has been selected for the Tracker.
- **Greece**: Greece only releases PDFs which contain information at the prefecture level. This information is downloaded from a GitHub repository that scrapes the PDFs. The data which is at the prefecture level, matches perfectly to NUTS 2 boundaries.
- **Hungary**: Hungary does not officially release any data. A map is uploaded in an image format on the official website which is scrapped daily. This information is retrieved from one of the several GitHub repositories that keep track of cases in Hungary.
- **Ireland**: Ireland releases regional data on ArcGIS Hub which is downloaded manually for processing. The links are provided in the Stata script.
- **Italy**: Italy has one of the best data-sharing setup among the European countries. Italy currently provides full documentation and access to their data on their official GitHub page (https://github.com/pcm-dpc/COVID-19). The regions defined in the data perfectly match NUTS 2021 definitions but the aggregation to NUTS 2016 has to be approximated for three small island regions since their boundaries were modified. See Technical Validation sector for details.
- **Latvia**: Latvia updates the data daily which can be downloaded directly from the official website. Regions with cases under 5 are displayed with a range of 1-5 for privacy reasons. This has been replaced with a value of one to keep the variable numeric. The raw file containing the original information can used to process these ranges differently.
- **Netherlands**: Netherlands data is downloaded directly from the official website for processing.
- **Norway**: Since Norway is outside of the EU, no consistent information at the NUTS 3 level is available. The Norwegian data is provided at the Kommune level, a tier below NUTS 3. This data is spatially joined with the NUTS 2016 boundaries to create a Kommune-to-NUTS 3 crosswalk which results in a perfect mapping. See the Technical Validation section for details.
- **Poland**: Poland data is extracted from a GitHub repository that is mapped perfectly to NUTS 2 boundaries. Poland launched its data sharing service relatively recently which currently also provides NUTS 3 level, but it does not have the temporal extent to fit in this Tracker. Minor data errors exist since part of the data is scraped. Please see the Technical Validation section for details.
- **Portugal**: The official regional data for Portugal has major issues. Data on regions was released on a daily frequency till July 2020, after which, it was switched to a weekly frequency. Currently information is only available on a bi-weekly basis and therefore there are major gaps in the daily cases data. Out of all the countries in the Tracker, Portugal’s data is currently the only one that is not at a daily frequency.
- **Romania**: The official data for Romania is available in JSON format which is converted into a CSV format for processing in Stata.
- **Slovak Republic**: The data for Slovak Republic is downloaded from a GitHub repository which processes the official data.
- **Slovenia**: The data for Slovenia is downloaded from a GitHub repository which processes the official data.
- **Spain**: Spain’s data is downloaded directly from the official website.
- **Sweden**: Sweden releases a file daily which is manually downloaded.
- **Switzerland**: Switzerland does not have country-wide information available in a centralized place. Instead, a group of independent researchers collate this information from the official websites of the various Cantons, which are also NUTS 3 regions. Therefore the very latest data is not immediately available.
- **United Kingdom**: The data for UK is not easily accessible. Despite shifting to a new website in the summer of 2020, information is only released for 7-day intervals for regions below NUTS 3 in the form of an interactive map (https://coronavirus.data.gov.uk/details/interactive-map). Furthermore, the data portal for the UK (https://coronavirus.data.gov.uk/details/download) has put significant restrictions on bulk downloading. Therefore, UK is dealt with as four separate countries; England, Scotland, North Ireland, and Wales. Daily regional data has been found only for the following two countries:
- **England**: In the early days of the pandemic an independent researcher, Tom White (https://github.com/tomwhite/covid-19-uk-data), put enormous amounts of effort to collate UK information from various sources. His work was later picked up by ODI Leeds (https://github.com/odileeds/) that regularly update the dataset for England. The data is available at the Local Authority Districts (LAD) level using April 2019 definitions. LADs aggregate perfectly to NUTS 3 2016 boundaries. See the Technical Validation section for details.
- **Scotland**: Data for Scotland is pulled directly from the Public Health of Scotland website (https://www.opendata.nhs.scot/dataset/covid-19-in-scotland). Scotland’s data is processed using the same routine as England where LAD April 2019 boundaries are matched to NUTS 3. See the section Technical Validation for details.

The following countries – Albania, Bosnia, Bulgaria, Serbia, Lithuania, North Ireland (UK), North Macedonia, Turkey, and Wales (UK), have official NUTS 3 correspondence tables but are not currently in the Tracker since their regional data has not been located. If these countries have to be used, their data can be replaced with country-level indicators to complete the map. For analysis this is still useful as cases normalized by population allow for comparison of regions of various sizes. As a note of caution, the above information and descriptions are subject to change as countries evolve their COVID-19 data sharing strategies. Please check the GitHub page (https://github.com/asjadnaqvi/COVID19-European-Regional-Tracker) for the latest updates.

### Overall data trends

Figure 3 shows the data points available for each country. Here one can note major gaps in Portugal, Greece, and Estonia daily cases data but on average the remaining countries are mostly complete. The data version used for this paper has over 500,000 data points. Individual NUTS-level cases per 10,000 population are plotted in Figure 4. In this figure we can also observe the difference in normalized daily new cases between the first and the second waves. The spread goes up significantly in the second wave.

**Figure 4.**
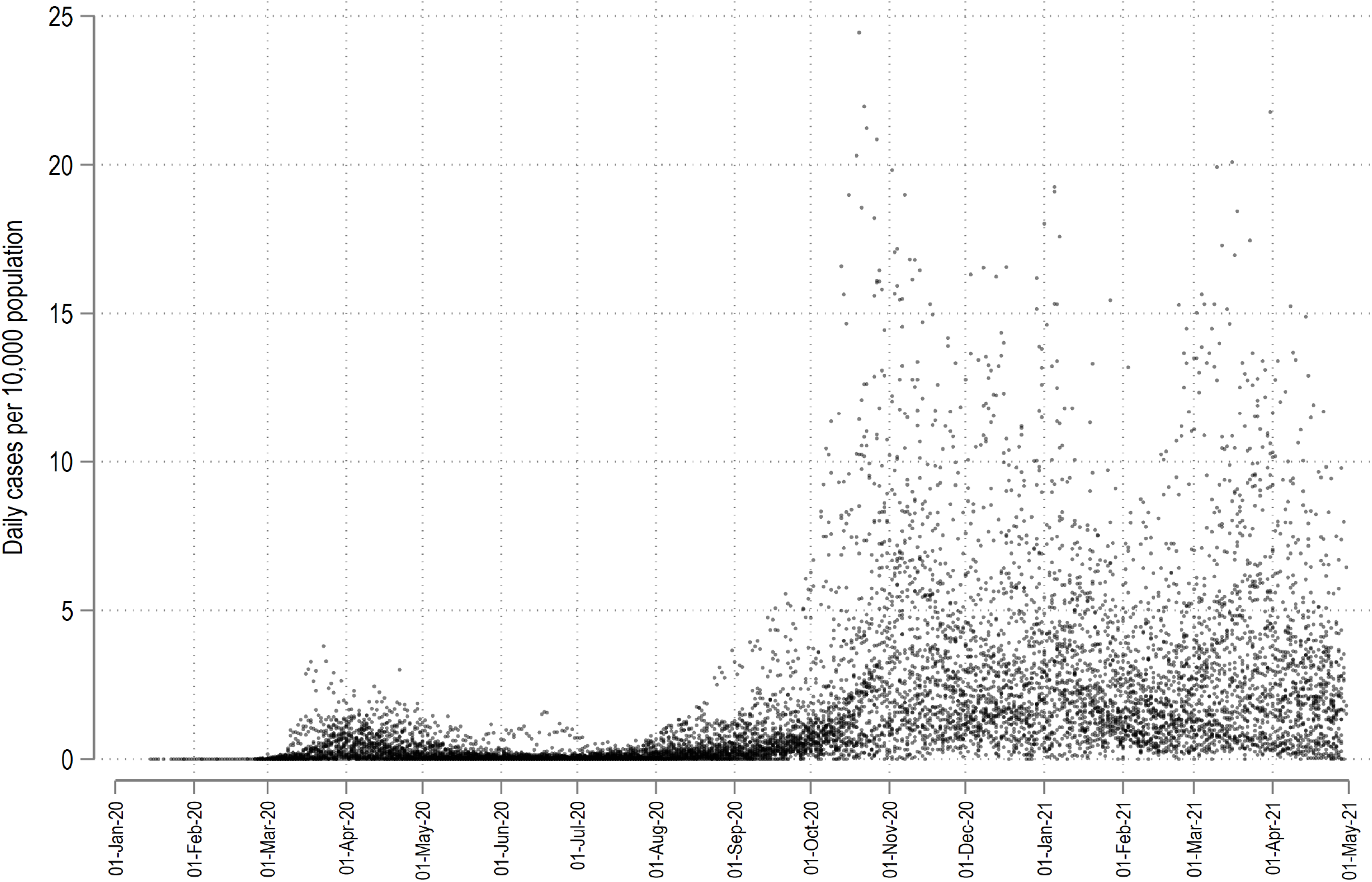
Distribution of daily cases per 10,000 population.

Figure 5 shows the cumulative distribution of cases and cases per 10,000 population for the data in this Tracker that ranges from January 2020 to April 2021. Here we can immediately observe that larger units in terms of area have more cases overall than smaller ones. If we control for population size then a different picture emerges. For example, it is clear from the cumulative cases per 10,000 population map that Germany managed to insulate itself very well from neighboring countries by enforcing strict border controls and lockdown policies. Despite this, regions on the east side show a slightly higher incidence rate than western and northern Germany. Similarly the western part of Austria, north Italy, Switzerland, and eastern part of France have a much higher cases per capita relative to the remaining regions within these countries. Sweden with its lack of strict lockdown policies also stands out among the Nordic countries. Explanations for these trends are left as research questions. Furthermore, given the daily resolution of the data in the Tracker, these maps can also be checked for variations in the first and the second waves.

**Figure 5.**
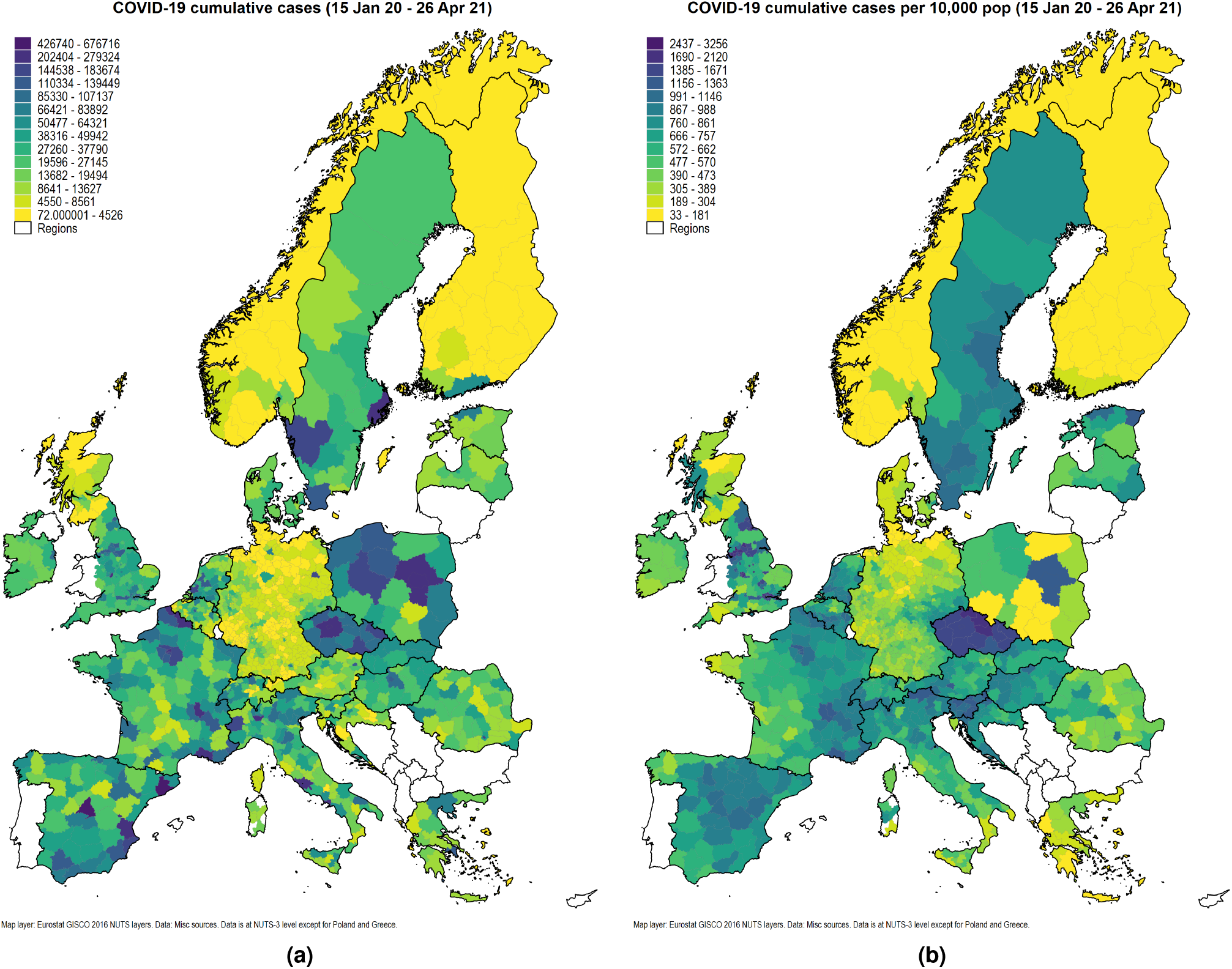
Spatial distribution of cumulative COVID-19 cases.

**Figure 6.**
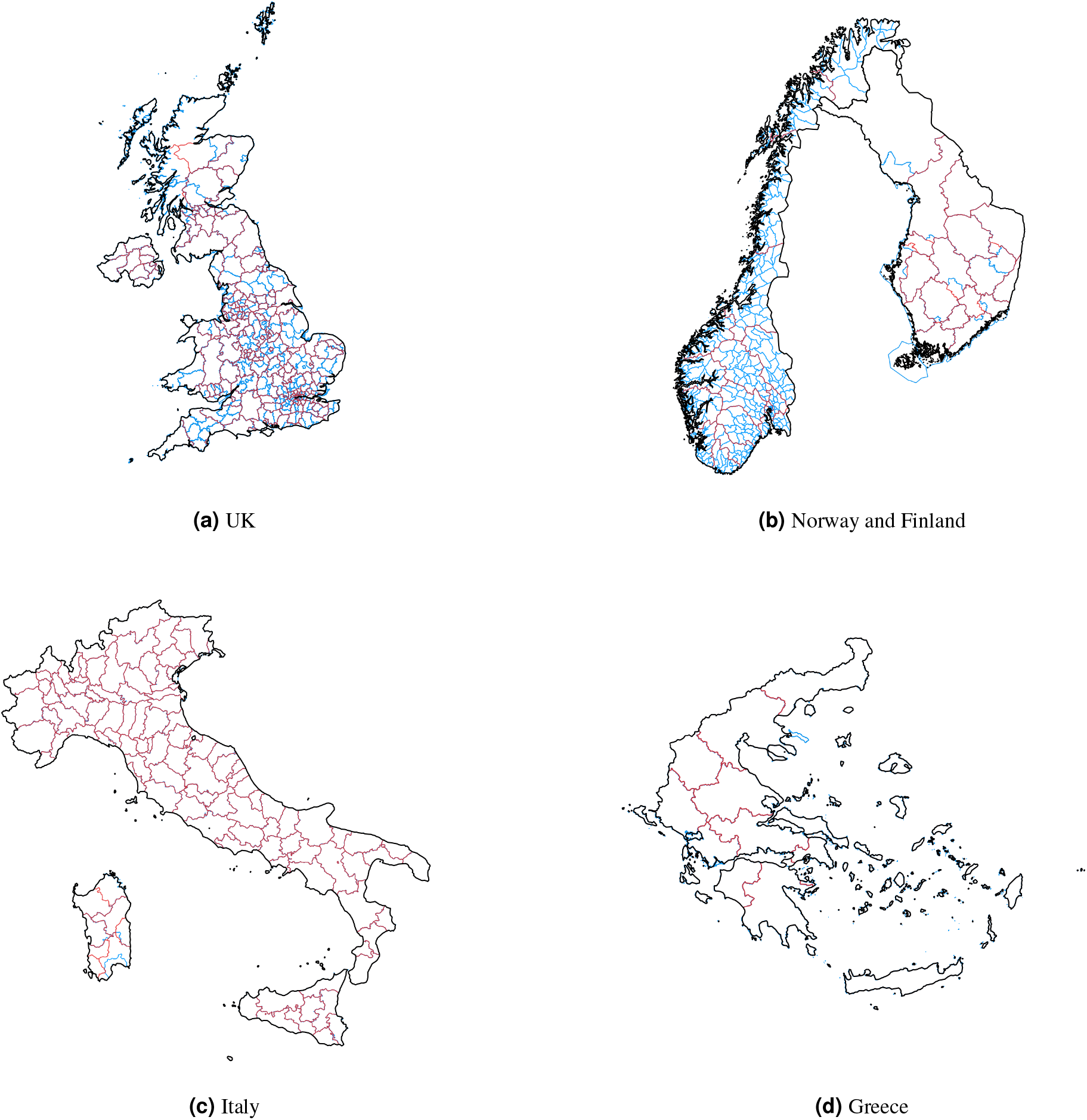
Countries where region-to-NUTS boundaries are approximated.

### Technical Validation

Three steps have been taken to ensure that veracity and accuracy of the information and to document errors that might occur in the homogenized file.

First, since the country-level datasets are official records provided by different government departments of each country, they can be compared with various online dashboards highlighted in Table 2. It is also important to point out that not all countries release the latest data at the regional level. A good example of this is France, that releases a file daily but the regional information is usually two to three days old. Similarly the crowd-sourced data for Switzerland, is back-filled data as information for the different regions (Cantons) is updated. Regardless of these lags, comparison of values with online dashboards is possible for most countries in the Tracker.

Second, as discussed earlier, regional data is approximated during the homogenization process for some countries. In order to ensure transparency, Table 3 provides notes for various countries where either data is converted from ranges into unique values (for example, Belgium, Estonia, and Latvia) or boundaries are approximated using a spatial merge (for example, Finland, UK, Norway, and Greece). The *data approximation* error occurs when ranges are converted into unique numerical values. This is a judgement call in order to allow these countries to merge with the remaining files. For Belgium and Latvia, regions with cases less than five are anonymized as *<*5. These values have been replaced with 1. For Estonia, which only provides data in ranges of 10s, mid values of each range is taken and multiplied with total cases to estimate cases per region, and then the regions are aggregated to NUT 3. Therefore, the data approximation is the highest for Estonia. The raw data and the scripts are available in case users prefer the actual information or have other approximation strategies.

**Table 3.**
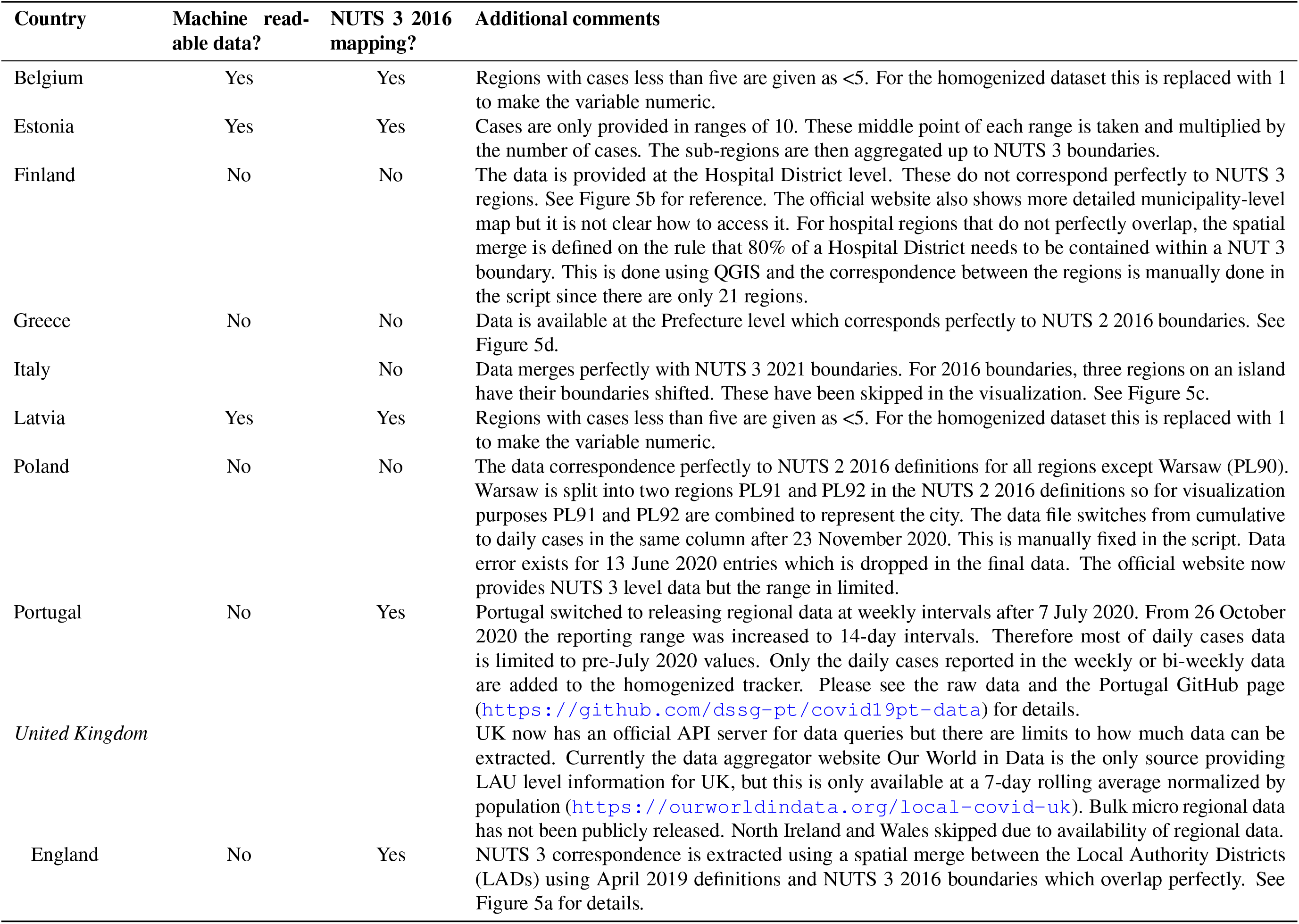
Countries with data or mapping issues

For countries which do not have an official correspondence to NUTS regions, a *boundary approximation* is done with a spatial merge by overlaying administrative boundaries with NUTS boundaries. For UK, Norway and Greece, the spatial merge creates a perfect mapping as shown in Figure 5. For Finland, the hospital districts, that are size-wise comparable to NUTS 3 boundaries contains minor errors in some cases since boundaries do not align. In an ideal case scenario, data should be provided at the smallest possible unit that can be aggregated up to various administrative boundaries, as is the case with UK and Norway. The official Finish website (see Table 2) does provide COVID-19 data at the municipality level, a unit below NUTS 3, but accessing the map is not straightforward nor it is clear whether this data can be accessed. On a related note, Italy data is officially released at the NUTS 2021 definitions so mapping to NUTS 2016 definitions do not work perfectly for three regions on an island off mainland Italy. These regions show up as missing on the map. For both the Finnish and Italian regions that do not perfectly match, the number of cases are very small which also keeps the error rate small. Regardless, original administrative units and their data in the raw files and can be used for higher accuracy.

Third, since the data is at the regional level, it can be aggregated up to generate country-level totals which can be compared with data aggregation websites like the Our World in Data (OWID) COVID-19 tracker (https://ourworldindata.org/coronavirus). OWID is used and referenced almost daily in scientific research and the media and has a major impact of policy discussions. OWID was utilizing country-level information provided by the European Center for Disease Control (ECDC) till November 2020. In November 2020, ECDC announced that it will switch to weekly data releases under The European Surveillance System (TESSy) (https://covid19-surveillance-report.ecdc.europa.eu/) where countries directly submit aggregated NUTS 2 level data. As a response OWID switched to the John Hopkins University’s (JHU) data repository (https://coronavirus.jhu.edu/map.html), a major data source for COVID-19 information at the global level.^16^ For validation, both this Tracker and OWID data is merged on a country-date combination and the difference between the daily cases is calculated. Figure 7 plots the difference split by countries and highlights how the match quality goes down after 1 October 2020. After October 2020, the mismatch for most countries increases significantly and persists till today. This highlights two points. First, before October 2020, data was provided by ECDC which was taking information directly from European countries. Since this Tracker is also pulling data from the countries directly, the match is very close with the exception of some outliers. This exercise helps validate the data of this Tracker. Second, since the data source of this Tracker remains the same, while OWID changed its source to a more unverified data set after October 2020, this Tracker provides a more accurate picture of country-level aggregates and also includes regional variation. As of March 2021, ECDC is again providing access to daily country level data but gaps exist between the new data series and the pre-November 2020 updates.^15^

**Figure 7.**
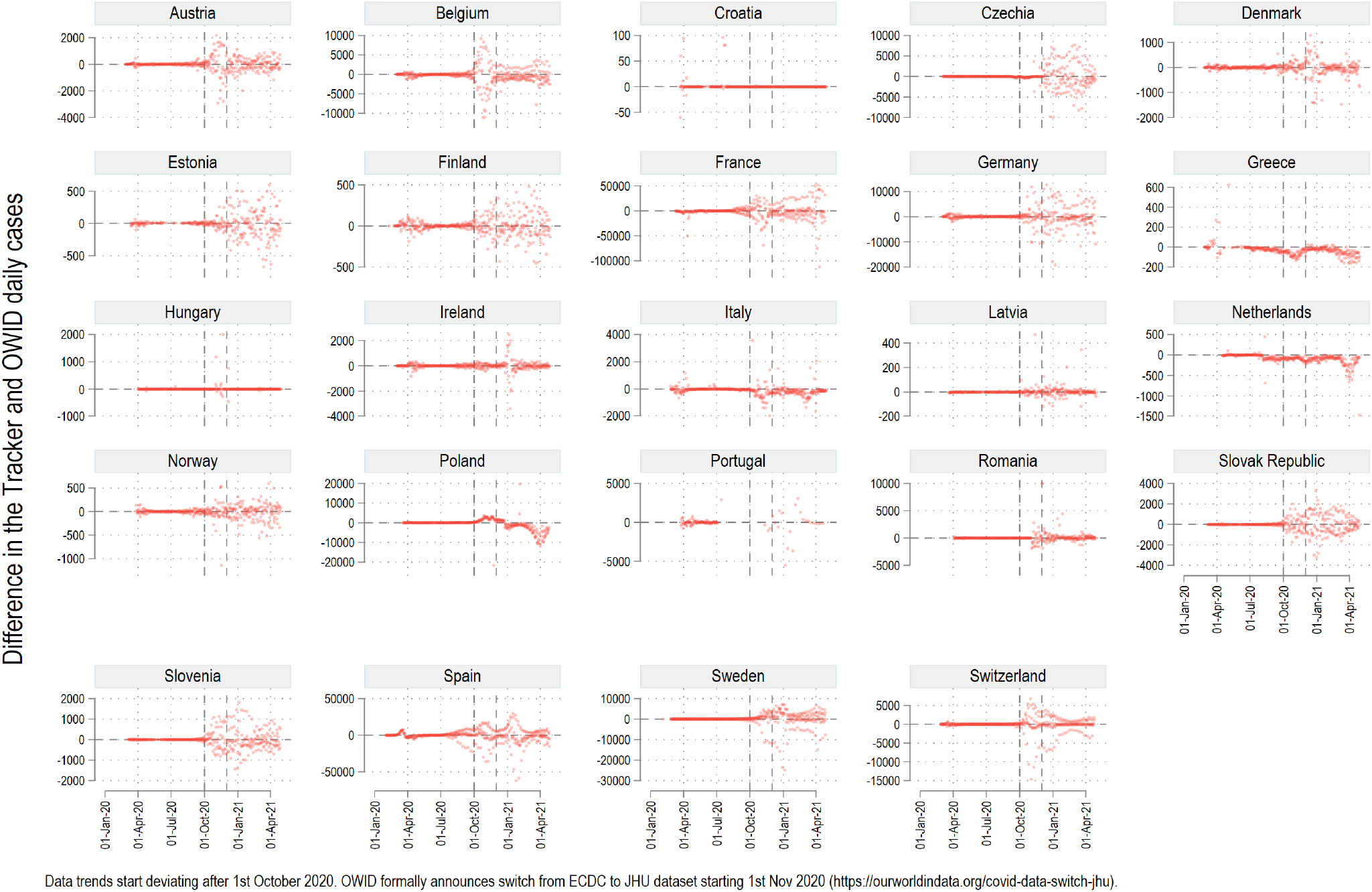
Tracker comparison with Our World in Data (OWID) values.

### Usage Notes

The dataset has been compiled in Stata (www.stata.com), a standard statistical software mostly used in the field of economics. All data including raw files, scripts, and the final data set are provided on GitHub (). Besides the Stata .dta data format, all information is also stored in the generic .csv format allowing the data to be accessed in any software. Annotated Stata scripts, or dofiles are also provided. Stata has an easy-to-interpret syntax structure that can be easily translated into other programming languages.

As a caveat, there are two features that Stata cannot handle well. First, is the ability to download files that are redirected from clicks or links on websites. This level of web-scraping works much better in other languages designed for such tasks like R or Python. Second data in JSON format has to be parsed either manually using user-written commands, or via third-party JSON-to-csv converters that are available online. Processing complex data structures like JSON or XML are also easier in other languages.

The repository is updated approximately every four weeks including a public release on Zenodo. If users prefer a higher frequency of updates, then the Stata scripts can be used directly to process the files or a more frequent release can be requested as well. Since the aim of the homogenized dataset is to be able to correlated it with existing NUTS level datasets, a monthly frequency is sufficient since European-level regional data takes several weeks or even months to be released.

This Tracker can be utilized for a host of different research directions. For example, the Tracker can be mapped onto NUTS-level regional data (https://ec.europa.eu/eurostat/web/regions/data/database) released by Eurostat.^3^ This includes various economic, demographic, health, tourism, and labor related indicators some of which also have a monthly or even a weekly frequency. Since data for individual countries is also provided, a detailed country-specific analysis can also be done provided regional or micro data is available for analysis. Other datasets catalogued on the Oxford COVID-19 Supertracker^13^ provides a range of interesting information on various policies put in place by countries during the pandemic. The Tracker data can also be combined with several innovative global datasets which contain NUTS-level information for European countries. This, for example, includes Google Mobility trends^4^ or the Facebook Social Connectivity Index.^5^ Since, the data for the Tracker has a Creative Commons Attribution 4.0 International Licence (CC-BY), anyone can access it at any point in time, and it will be regularly updated until the countries stop publishing regional COVID-19 data.

## Code Availability

The latest code to process the files can be downloaded from the Zenodo release^1^ or accessed from the GitHub repository (https://github.com/asjadnaqvi/COVID19-European-Regional-Tracker). The code is written in Stata versions 15 and higher and these are also recommended for improved functionality with maps and graphs. Data processing can be done in any version.

As specified in the Data Records section, the dofiles are in the **/02 dofiles/** folder. Within this folder dofiles exist for each country plus a set of five dofiles that setup, merge, map, and validate the final data file. These files are explained as follows:

- COUNTRY_SETUP.do initializes the code for running the country files. One can run each country file independently as well, but they need the directory structure and packages to be loaded in order to function correctly. Directory and packages can be initialized using the first few lines marked in the beginning of the COUNTRY_SETUP.do file. This syntax is as follows:

~~~
clear
global coviddir “<your directory path>/<your directory name>“
* package for maps and correcting map projections
ssc install spmap, replace
ssc install geo2xy, replace
* packages for color schemes
ssc install palettes, replace
ssc install colrspace, replace
* package for replicating plots
net install tsg_schemes, ///
from(“https://raw.githubusercontent.com/asjadnaqvi/Stata-schemes/main/schemes/“)
set scheme white_w3d, perm
set the detault graph font
graph set window fontface “Arial Narrow”
~~~

Each country .do file is annotated with notes where necessary.

- COUNTRY_MERGE.do combines all the country datasets saved in **04_master** in one file EUROPE_COVID19 master.dta. The master file is also saved in the **04_master** folder.
- COUNTRY_GIS_setup.do sets up the GIS layers in Stata format for the combined NUTS regions and for individual countries. A mixed NUTS3 and NUTS2 shapefile is also created accommodate the data from Poland and Greece. The logic can also be applied to add data at the provincial (NUTS 1) or country (NUTS 0) level if one needs to add other countries not in the dataset. This file also extracts shapefiles for individual countries and generates a file used for labeling the individual country maps.
- COUNTRY_GIS_map.do create the maps that are saved in the **05_figures** folder. See Figure 5 for the overall map. Individual country COVID-19 maps can be viewed on GitHub (https://github.com/asjadnaqvi/COVID19-European-Regional-Tracker).
- COUNTRY_validation.do collapses the Tracker to a country-date level and merges it with OWID COVID-19 dataset, for validation. This file also produces Figure 7.

GitHub and Zenodo files are updated every four weeks. Each Zenodo release is assigned a unique DOI, but the generic DOI: https://doi.org/10.5281/zenodo.4244878, will always link to the latest version.^1^

## Data Availability

The data for the paper has been made publicly available on Zenodo under the Creative Commons Attribution 4.0 International Licence (CC-BY)

https://doi.org/10.5281/zenodo.4244878

## Acknowledgements

I would like to thank the International Institute for Applied Systems Analysis’ (IIASA) directors, Albert van Jaarsveld, and Leena Srivastava, and the Director of the Advancing Systems Analysis (ASA) program, Elena Rovenskaya, for their encouragement and continuous support throughout the course of this Tracker. I would also like to thank IIASA for partially funding this project.

## Author contributions statement

A.N. set up the Tracker, including identifying the websites and the relevant files, defining the protocols for the workflow and data management, writing the code, and cleaning up the data. A.N. will be responsible for updating the Tracker on a monthly basis till the countries stop reporting their data.

## References

1. Naqvi, A. COVID-19 European regional tracker. Zenodo 10.5281/zenodo.4244878 (2021).

2. Eurostat. Nomenclature of Territorial Units for Statistics (NUTS). Website https://ec.europa.eu/eurostat/web/nuts/ background (2021).

3. Eurostat. European Statistical Agency. Website https://ourworldindata.org/coronavirus (2021).

4. Google. Community mobility reports. Website https://www.google.com/covid19/mobility/ (2021).

5. Facebook. Data for good: Social Connectedness Index. Website https://dataforgood.fb.com/tools/ social-connectedness-index/ (2021).

6. Eurostat. ECDC statement on the rapid increase of COVID-19 cases in Italy. Newspaper article https://www.ecdc.europa. eu/en/news-events/ecdc-statement-rapid-increase-covid-19-cases-italy (23 Feb 2020).

7. Eurostat. Coronavirus: Ischgl resort at heart of Europe’s outbreak reopens. Newspaper article https://www.bbc.com/news/ world-europe-52384572 (12 Oct 2020).

8. Deutsche Welle (DW). Coronavirus: What are the lockdown measures across Europe? Newspaper article https://www.dw.com/en/coronavirus-what-are-the-lockdown-measures-across-europe/a-52905137 (14 April 2020).

9. Euronews. How hard is the coronavirus second wave hitting in Europe?. Newspaper article https://www.euronews.com/2020/11/13/europe-s-second-wave-of-coronavirus-here-s-what-s-happening-across-the-continent (13 November 2020).

10. Oxford. Oxford COVID-19 Government Response Tracker (OxCGRT). Website https://www.bsg.ox.ac.uk/research/research-projects/coronavirus-government-response-tracker (2021).

11. Desvars-Larrive, A. et al. A structured open dataset of government interventions in response to COVID-19. Sci. Data 7, 285, 10.1038/s41597-020-00609-9 (2020).

12. Hasell, J. et al. A cross-country database of COVID-19 testing. Sci. Data 7, 345, 10.1038/s41597-020-00688-8 (2020).

13. Oxford. Oxford Supertracker: The global directory for COVID policy trackers and surveys. Website https://www.bsg.ox.ac.uk/research/research-projects/coronavirus-government-response-tracker (2021).

14. ECDC. COVID-19 database European Centre for Disease Prevention and Control. Website https://www.ecdc.europa.eu/en/covid-19/data (2021).

15. ECDC. COVID-19 situation update for the EU and the EEA. Website https://www.ecdc.europa.eu/en/cases-2019-ncov-eueea (2021).

16. Our World in Data. Our World in Data switches to Johns Hopkins University as our main data source for COVID-19 cases and deaths. Online post https://ourworldindata.org/covid-data-switch-jhu (2020).

17. Eurostat. Local Administrative Units (LAUs). Website https://ec.europa.eu/eurostat/web/nuts/local-administrative-units (2021).

18. GISCO. The Geographic Information System of the Commission - NUTS Statistical Administrative Units Shapefiles. Website https://ec.europa.eu/eurostat/web/gisco/geodata/reference-data/administrative-units-statistical-units/nuts (2021).

